# Environmental modifiers of developmental outcomes in genetic epilepsy

**DOI:** 10.64898/2025.12.03.25341548

**Authors:** Christian M. Boßelmann, Natasha N. Ludwig, Calliope Holingue, Andres Jimenez-Gomez, Andrea Ganna, M Scott Perry, Ana Arenivas, Dennis Lal

## Abstract

Genetic neurodevelopmental disorders (NDDs) with epilepsy are genome-defined yet exposure-sensitive. While the mechanistic effects of genetic variants underlying these disorders are widely studied to understand clinical heterogeneity among patients, the contribution of external environmental factors remains largely unexplored. Using the Simons Searchlight population dataset, we examined caregiver-reported environmental exposures and functional outcomes from 970 individuals with (67%) and without (33%) epilepsy across 93 NDDs. The primary outcome was adaptive function (Vineland-3), with quality of life (QI-Disability) and autism symptoms (SCQ Lifetime) as secondary analyses. Compared to individuals with NDD without epilepsy, individuals with epilepsy were more likely to be admitted for inpatient or intensive care and more likely to receive treatment with sodium channel blockers; however other exposures were similar across groups. While quality of life was similar, those with epilepsy had lower adaptive function (*p* < 0.001). On multivariable regression analysis, we found positive associations between socioeconomic factors and quality of life (parents’ highest educational status: *β* = 0.014, 95% CI: [0.003, 0.025], *p* = 0.013) and annual household income: *β* = 0.007, 95% CI: [0.001, 0.014], *p* = 0.021) and negative associations between treatment-related factors and adaptive function (treatment with sodium channel blockers: *β* = −0.087, 95% CI: [−0.127, −0.047], *p* = 2.44×10^-5^; hospitalisations: *β* = - 0.055, 95% CI: [−0.076, −0.034], *p* = 3.65×10^-7^), among others. These associations were mediated by epilepsy duration and severity, burden of polytherapy, and the underlying genetic aetiology, suggesting that genetics-informed care and early seizure control may facilitate better developmental outcomes. Results were consistent across subgroups and sensitivity analyses but were not observed in comparison groups of 322 and 1,276 individuals without epilepsy, suggesting gene-, disease-, or treatment-specific effects in epilepsy. Together, environmental exposures explained significantly more variance in developmental outcomes than genetic aetiology alone (additional *R*^2^ = 19.6%, *p* = 0.003). Importantly, the additional variance explained by environmental exposures was different across genes and ranged from ∼15% (*SCN2A, SLC6A1*) to >30% (16p11.2 deletion), suggesting gene-specific vulnerability to external factors. Overall, 38-58% of variance in developmental outcomes remained unexplained, supporting further data collection and the identification of additional genome and exposome influences. Collectively, these findings reframe genetic NDDs as genome-informed yet exposure-sensitive disorders and point to early, genetically informed seizure management and careful stewardship of treatment exposures as immediate levers to improve developmental trajectories.

## Introduction

Neurodevelopmental disorders (NDD) are heterogeneous conditions including attention deficit hyperactivity disorder (ADHD), autism spectrum disorder (ASD), developmental delay (DD), intellectual disability (ID), and other disorders of communication, learning, and motor skills.^1,2^ There is a considerable clinical overlap with epilepsy, often as developmental and epileptic encephalopathies (DEE) characterized by both treatment-resistant seizures and developmental plateau or regression.^3,4^ This overlap relates to the shared genetic background of NDD and DEE: Both groups of disorders may be caused by disruptions of critical brain-specific mechanisms including neuronal differentiation, synaptic or ion channel function, and neurotransmitter signaling.^5–7^ Beyond the genetic aetiology, there has been an increasing awareness of external environmental factors in these groups of disorders.^8,9^ These environmental factors may potentially be modifiable, allowing for improved outcomes in these often severely affected individuals. This directly represents a priority objective of the World Health Organization (WHO), which has identified epilepsy as one of the most common underlying and avoidable causes of mortality in individuals with NDD.^10^

Individuals with NDDs with epilepsy are subject to considerable additional disease burden from early exposure to multiple anti-seizure medications (ASMs), frequent seizure-related hospitalisations including intensive care, and higher risk of seizure-related death (SUDEP). These factors are not typically present in NDDs without seizures.^11,12^ We have previously shown that individuals with NDDs with epilepsy are hospitalized almost twice as often as those without epilepsy, especially during developmentally important periods including early childhood and transition to adulthood.^12^ They are also more likely to receive broad-spectrum and syndrome-specific ASMs with potentially severe side effects.^12^ This observation of potentially modifiable factors in a vulnerable subgroup – individuals with NDDs with epilepsy – motivated our analysis of developmental outcomes. Prior studies investigating developmental outcomes of NDD with epilepsy were generally limited to small and gene-specific cohorts.^13,14^

Here, we leveraged insights from 970 individuals in the Simons Searchlight population dataset by the Simons Foundation Autism Research Initiative to investigate environmental and genetic determinants of developmental outcomes across a comprehensive spectrum of 93 NDDs with and without epilepsy. For this exploratory study, we investigated the following exposures based on previous literature: *(i)* pregnancy-related factors (maternal use of alcohol or tobacco; supplementation of folic acid, including prenatal vitamins, or iron during pregnancy; preterm birth)^15–18^; *(ii)* family-related and socioeconomic factors (parents’ marital status; educational status; household income; foster care)^19–21^; and *(iii)* treatment-related factors (exposure to sodium channel blockers (SCBs); admission to inpatient or intensive care).^22–25^ Importantly, we used caregiver-reported measures of development and quality of life as outcomes. While conferring potential biases outlined below, caregiver-reported outcomes offer a comprehensive view of a child’s experience and focus on individually meaningful aspects of living.

## Materials and methods

### Study Population

This study was based on the Simons Searchlight population dataset (v13.0, release 2025–06-24) by the Simons Foundation Autism Research Initiative (SFARI). Briefly, the Simons Searchlight population dataset includes families of individuals with any of 24 copy-number variants and 160 monogenic disorders associated with autism and NDD as well as their families, of which 93 genetic etiologies were represented in our study cohort (Supplementary Methods; Supplementary Table 1). Inclusion criteria for our analysis were: *(i)* affected individuals (initially identified probands) with any genetic NDD; *(ii)* at least one score assessment for the primary outcome, the Vineland Adaptive Behaviour Scales, Third Edition. The remaining 2,246 individuals were split into three groups: *(i)* the main analysis group who had a diagnosis of epilepsy (*n =* 648); *(ii)* a comparison group of affected individuals with any genetic NDD who did not have a diagnosis of epilepsy (*n =* 322), used for comparison of baseline clinical and demographic characteristics and investigation of epilepsy-specific outcomes; *(iii)* individuals where information about epilepsy was not available (*n =* 1,276), which included the previous group and was used for sensitivity analyses only (Fig. 1). Data are reported in line with the Strengthening Reporting of Observational Studies in Epidemiology (STROBE) statement.

**Figure 1.**
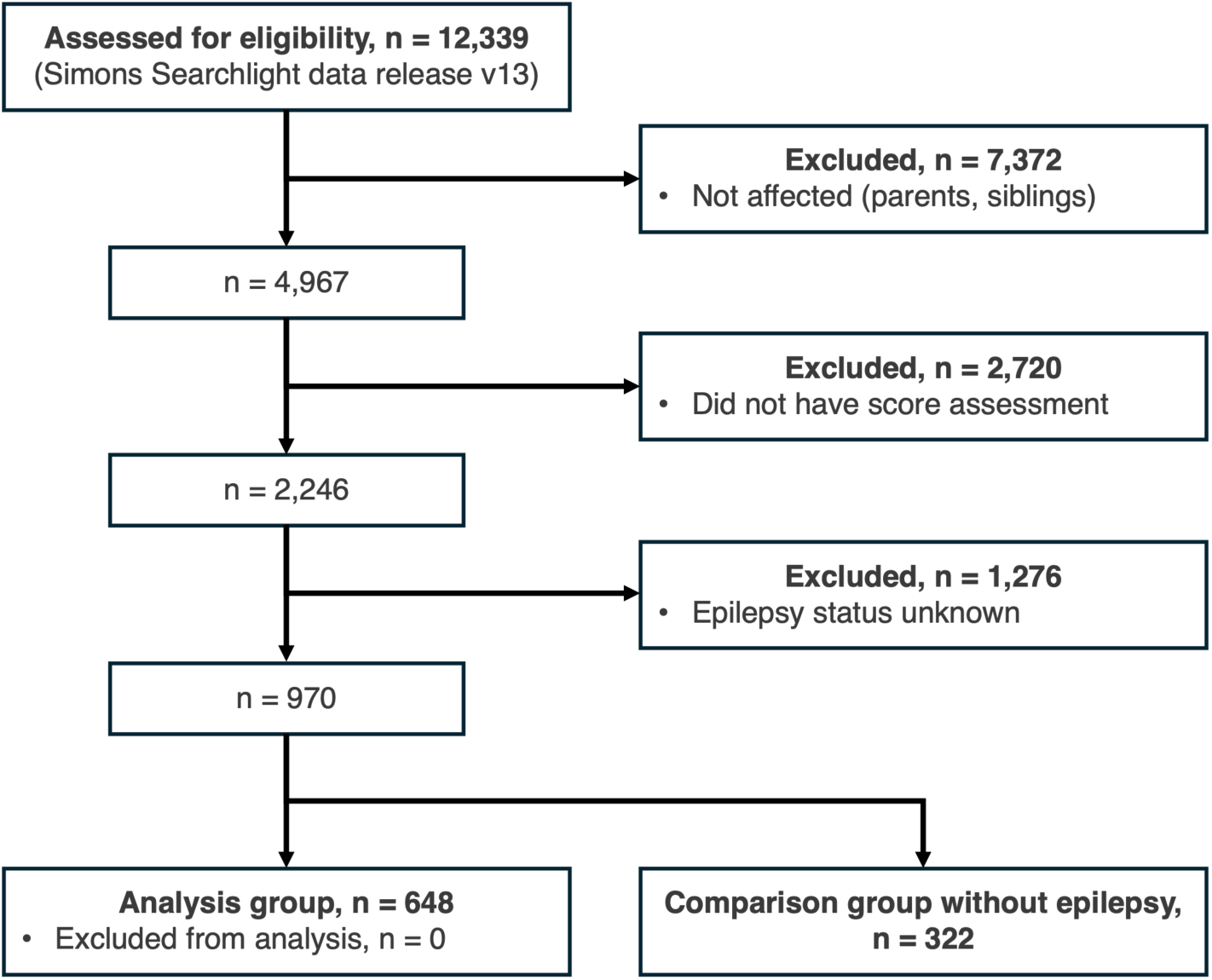
CONSORT flow diagram.

### Measurements

Predictors, (i.e., environmental factors including socioeconomic and treatment-related factors hypothesized to be associated with the included outcomes) were selected based on data availability (presence in the Simons Searchlight population dataset) and previous literature. A full list of available and selected measures, including the rationale for selection and count by genetic aetiology, is provided in Supplementary Table 1. These hypotheses were selected to reduce the likelihood of spurious false-positive associations given that Simons Searchlight offers in-depth phenotypic data. These predictors included: *(i)* pregnancy-related factors (exposure to alcohol, exposure to tobacco, folic acid supplementation, iron supplementation, preterm birth); *(ii)* family-related and socioeconomic factors (parents’ marital status, parents’ highest educational status, annual household income, foster care or home placement); and *(iii)* treatment-related factors (any ICU admission, number of hospitalisations, exposure to SCBs – including lamotrigine, carbamazepine, oxcarbazepine, eslicarbazepine, phenytoin, lacosamide). Dosage information for SCBs was not available. Prescriptions for other ASMs were used in further subgroup and sensitivity analyses.

Developmental outcomes were investigated using three questionnaires, which were routinely administered for parents or caregivers of individuals in the Simons Searchlight population dataset. We focused on outcome measures that were broadly available (>40%) to enable robust statistical analysis. For each outcome, the score at last follow-up was used. First, the Vineland Adaptive Behaviour Scales, Third Edition (Vinealnd-3) is a norm-referenced measure of adaptive function, a component of the DSM-V diagnostic criteria for ID.^2,26^ The Vineland-3 assesses functioning across three core domains (i.e., Communication, Daily Living Skills, and Socialization) yielding an Adaptive Behaviour Composite (ABC). There is also a Gross Motor Domain for those 9 years old and younger. The Vineland-3 yields standard scores (M = 100, SD = 15) and higher scores indicate greater adaptive function. The Vineland-3 has been used to measure non-seizure outcomes in rare genetic epilepsies in previous studies.^14,27^ Second, the Quality of Life Inventory-Disability (QI-Disability) was used to measure of quality of life in individuals with ID^28^. The QI-Disability yields a Total score and six domain scores (i.e., Health and Well-being, Positive Emotions, Negative Emotions, Social Interaction, Recreational Activities and the Outdoors, and Independence). Higher scores mean higher quality of life. The QI-Disability has previously been used in NDD with epilepsy related to variants in *CDKL5* and *SCN2A*.^13,29^ Third, the Social Communication Questionnaire (SCQ) Lifetime version score was used. This measure was developed to screen for symptoms associated with ASD and higher scores indicate greater likelihood of an ASD diagnosis.^30^ For the SCQ, three different thresholds based on previous sensitivity analyses were applied: ≥11 points (low), ≥15 points (standard), and ≥20 points (high).^31,32^ Raw scores from outcome measures were normalized at the gene level, using both min-max and z-score normalization, to account for gene-specific differences and enable cross-gene comparisons across the entire cohort. Raw scores were retained for sensitivity analyses.

### Statistical Analysis

We tested for associations between outcomes and predictors with multivariable linear (Vineland-3, QI-Disability) or logistic (SCQ Lifetime) regression models. Covariates for the main analyses were sex, ethnicity, and age at seizure onset, based on our previous study.^12^ Covariates for the sensitivity analyses are stated separately. Missing data were not imputed, and only complete cases were used for each model. Results were reported as effect estimates (beta) with 95% confidence intervals. Causal mediation effects were estimated using the mediate package at default parameters using 1000 simulations and quasi-Bayesian approximation. Continuous values across groups were compared using two-sided Wilcoxon rank-sum tests. Categorical values across groups were compared using Fisher’s exact test. Correlations between continuous values were reported with Pearson’s correlation coefficient. The nominal significance threshold was set at *α* = 0.05. Bonferroni correction for multiple testing was reported where appropriate. This study was carried out in the R programming language, version 4.4.2, with RStudio, version 2024.9.0.375.

### Data availability

The authors confirm that the data supporting the findings of this study are available within the article and its supplementary material. Approved researchers can obtain the Simons Searchlight population dataset described in this study (v13.0, release 2025–06-24) by applying at https://base.sfari.org.

## Results

### Environmental factors are associated with development in genetic epilepsy

Here, we report the relationship between environmental and treatment-related exposures and developmental outcomes across 970 individuals across 93 NDD. Most individuals had epilepsy (648/970, 66.8%) indicating that this represents both a common and a profoundly affected subgroup. The clinical and demographic characteristics of our study cohort are shown in Table 1. As expected, the distribution of genetic etiologies was different across groups: Monogenic disorders including channelopathies and synaptopathies were more common in the study cohort while deletion syndromes were more common in the comparison group of individuals in NDD without epilepsy. Quality of life was not significantly different between groups, but adaptive function was more impaired in individuals with epilepsy (*p* < 0.001). In total, 38/648 (5.9%) of individuals had the minimum score on the Vineland-3 scale, suggesting negligible bias from floor effects. The primary endpoint, Vineland-3, and secondary scores, QI-Disability and SCQ Lifetime, were moderately correlated (Pearson’s *r* 0.41-0.57, p < 0.001; Supplementary Fig. 1) while each score covered distinct functional domains. Scores were reported by caregivers at a median patient age of 10 years (IQR 6-16 years; Supplementary Fig. 2) and thus covered the relevant developmental period. After correcting for multiple testing, pregnancy-related factors and socioeconomic factors were not significantly different between groups. Individuals with epilepsy were more likely to be admitted to the intensive care unit (ICU), were more often hospitalised, and were more likely to receive treatment with sodium channel blockers (*p* < 0.001).

**Table 1.** Clinical and demographic characteristics of the study cohort. ^a^In the cohort of individuals without epilepsy, shown for comparison, 143/322 (44.4%) of individuals had at least one seizure, of which 38/322 (11.8%) also had received at least one ASM, but self-reported no formal diagnosis of epilepsy. Two-sided Wilcoxon rank-sum test for continuous variables and Fisher’s exact test for categorical variables. Genetic aetiologies are shown if they account for >5% of diagnoses in either group. *Significant after Bonferroni correction for multiple testing.

| | Study cohort (individuals with epilepsy, $n = 648$) | Comparison group (individuals without epilepsy, $n = 322$) | p-value |
| --- | --- | --- | --- |
| **Sex** |  |  |  |
| Male | 337 (52.0%) | 178 (55.3%) | 0.371 |
| Female | 311 (48.0%) | 144 (44.7%) |  |
| **Race** |  |  | 0.061 |
| White | 532 (83.1%) | 249 (78.8%) |  |
| More than one race | 66 (10.3%) | 44 (13.9%) |  |
| Asian | 18 (2.8%) | 16 (5.1%) |  |
| Other | 24 (3.8%) | 7 (2.2%) |  |
| **Genetic aetiology** |  |  | **<0.001\*** |
| SCN2A | 66 (10.2%) | 7 (2.2%) |  |
| STXBP1 | 60 (9.3%) | 9 (2.8%) |  |
| SLC6A1 | 55 (8.5%) | 10 (3.1%) |  |
| PPP2R5D | 41 (6.3%) | 27 (8.4%) |  |

**Table 1. Clinical and demographic characteristics of the study cohort.**
|  | Study cohort (individuals with epilepsy, n = 648) | Comparison group (individuals without epilepsy, n = 322) | p-value |
| --- | --- | --- | --- |
| *SYNGAP1* | 42 (6.5%) | 6 (1.9%) |  |
| *GRIN2B* | 14 (2.2%) | 25 (7.8%) |  |
| 16p11.2 deletion | 33 (5.1%) | 37 (11.5%) |  |
| Other aetiologies | 337 (52.0%) | 201 (62.3%) |  |
| **Clinical characteristics** |  |  |  |
| Age at first seizure (years), mean (SD) | 3.3 (3.8) | 4.0 (4.6)<sup>a</sup> | 0.030 |
| QI-Disability Total score, mean (SD) | 68.2 (15.1) | 70.1 (15.4) | 0.088 |
| SCQ Lifetime score, mean (SD) | 18.5 (7.7) | 17.1 (7.9) | 0.026 |
| SCQ Lifetime: non-verbal, n (%) | 221 (50.5%) | 136 (58.6%) | 0.053 |
| Vineland-3 ABC score, mean (SD) | 54.5 (21.2) | 61.0 (20.0) | **<0.001\*** |
| Vineland-3 ABC score 20, n (%) | 38 (5.9%) | 13 (4.0%) | 0.295 |
| Age at score evaluation (years), mean (SD) | 11.2 (7.8) | 10.6 (7.2) | 0.258 |
| **Treatment-related factors** |  |  |  |
| Any ICU admission, n (%) | 116 (18.0%) | 18 (6.5%) | **<0.001\*** |
| Number of hospitalisations, mean (SD) | 1.9 (1.0) | 1.2 (0.5) | **<0.001\*** |
| Sodium channel blocker treatment, n (%) | 317 (48.9%) | 17 (5.2%)<sup>a</sup> | **<0.001\*** |
| **Pregnancy-related factors** |  |  |  |
| Alcohol use, n (%) | 13 (2.0%) | 6 (1.9%) | 1.000 |
| Tobacco use, n (%) | 14 (2.2%) | 6 (1.9%) | 0.935 |
| Iron supplementation, n (%) | 22 (3.4%) | 10 (3.1%) | 0.993 |
| Folic acid supplementation, n (%) | 188 (29.0%) | 91 (28.3%) | 0.453 |
| **Familial and Socioeconomic factors** |  |  |  |
| Married biological parents, n (%) | 445 (71.5%) | 222 (71.6%) | 1.000 |
| Foster care, n (%) | 42 (6.7%) | 24 (7.7%) | 0.689 |
| Annual household income, n (%) |  |  | 0.804 |
| • Over \$161,000 | 117 (19.6%) | 50 (16.7%) | |
| • \$131,000 - \$160,000 | 49 (8.2%) | 27 (9.0%) | |
| • \$101,000 - \$130,000 | 78 (13.3%) | 46 (15.3%) | |
| • \$81,000 - \$100,000 | 79 (13.2%) | 37 (12.3%) | |
| • \$66,000 - \$80,000 | 47 (7.9%) | 22 (7.3%) | |
| • \$51,000 - \$65,000 | 64 (10.7%) | 33 (11.0%) | |
| • \$36,000 - \$50,000 | 64 (10.7%) | 31 (10.3%) | |
| • \$21,000 - \$35,000 | 60 (10.1%) | 26 (8.7%) | |
| • Less than \$20,000 | 39 (6.5%) | 28 (9.3%) | |
| Parents' Highest Education, n (%) |  |  | 0.014 |
| • Graduate/professional degree | 292 (47.4%) | 114 (37.3%) |  |
| • Baccalaureate degree (4-year degree) | 163 (26.5%) | 79 (25.8%) |  |
| • Associate degree (2-year degree) | 53 (8.6%) | 33 (10.8%) |  |
| • Some college | 64 (10.4%) | 54 (17.6%) |  |
| • GED diploma | 11 (1.8%) | 4 (1.3%) |  |
| • High school graduate | 27 (4.4%) | 18 (5.9%) |  |
| • Some high school (without diploma) | 6 (1.0%) | 3 (1.0%) |  |
| • Completed less than ninth grade | 0 (0.0%) | 1 (0.3%) |  |

For our main analysis, we investigated the associations between environmental factors and developmental outcomes using multivariable regression models with sex, race, and age at seizure onset as covariates. We first focused on our study cohort with a within-group analysis of individuals with genetic NDDs with epilepsy, while comparisons to the group without epilepsy and across genes are shown below. Outcomes were normalized for each gene, allowing for cross-cohort comparisons: For example, an effect estimate (*β*) of −0.2 corresponds to a 20% decrease for each one-unit increase in the predictor, across the range of developmental outcomes for each gene. We strictly selected twelve pre-specified hypotheses to reduce the likelihood of false-positive associations (Methods). Across these hypotheses, we found seven nominally significant associations. The strongest modifiers were treatment-related factors which were negatively associated with quality of life (treatment with SCBs: *β* = −0.034, 95% CI: [−0.064, −0.003], *p* = 0.031; hospitalisations: *β* = −0.026, 95% CI: [−0.042, −0.010], *p* = 0.002). Similarly, treatment-related factors were negatively associated with adaptive function (treatment with SCBs: *β* = −0.087, 95% CI: [−0.127, −0.047], *p* = 2.44×10^-5^; hospitalisations: *β* = −0.055, 95% CI: [−0.076, −0.034], *p* = 3.65×10^-7^; Fig. 2). Meanwhile, socioeconomic factors were positively associated with quality of life (parents’ highest educational status: *β* = 0.014, 95% CI: [0.003, 0.025], *p* = 0.013) and annual household income: *β* = 0.007, 95% CI: [0.001, 0.014], *p* = 0.021; Fig. 2).

**Figure 2.**
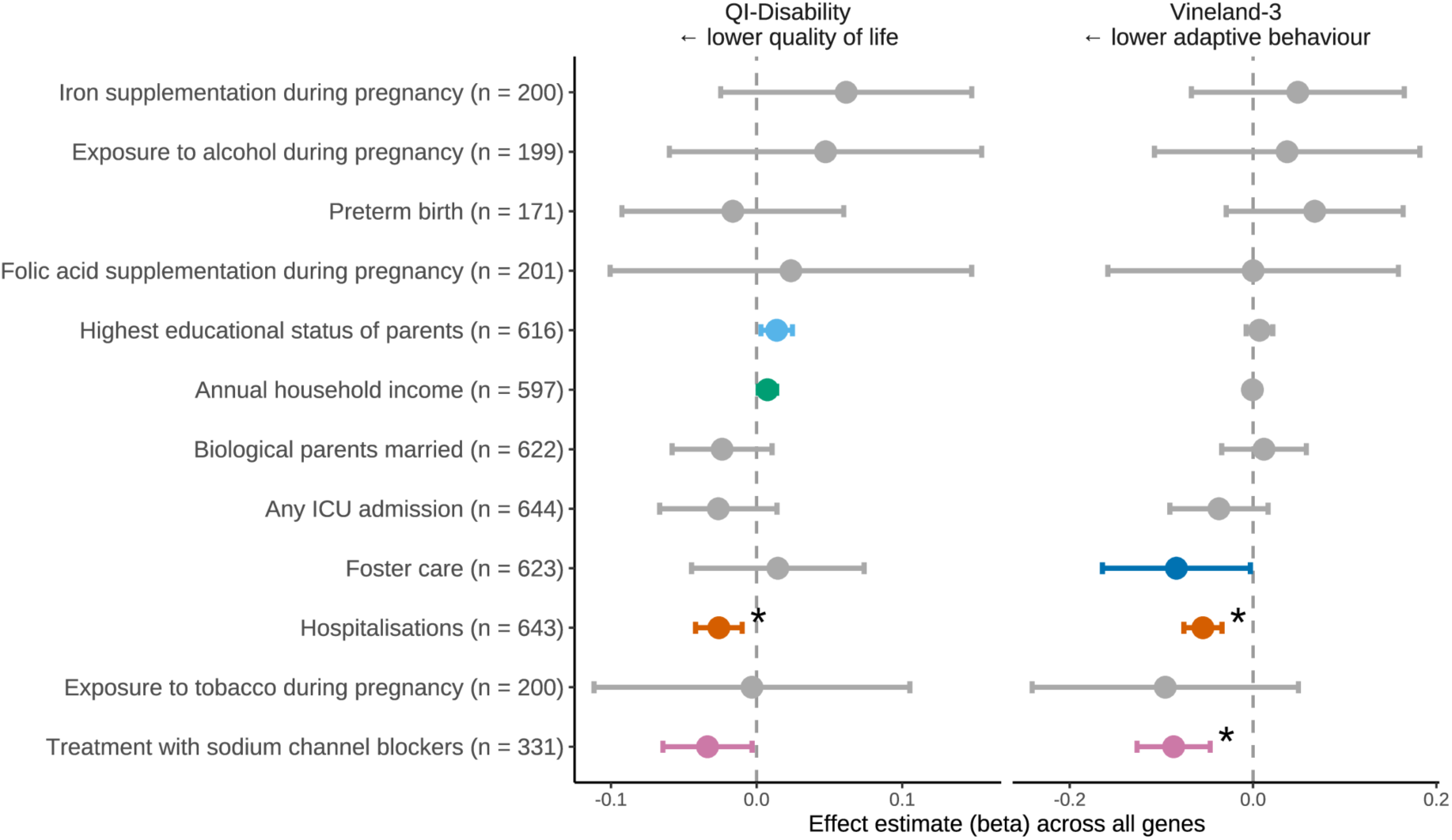
Environmental factors associated with developmental outcomes across genetic epilepsy syndromes. Forest plot of the effect estimates (beta) of exposure on gene-normalized Vineland-3 and QI-Disability scores. Multivariable linear regression with sex, ethnicity, and age at seizure onset as additional covariates. For each variable, the sample size, i.e. the number of individuals for which data was available in the Simons Searchlight population dataset (v13.0, release 2025-06-24), is shown in brackets. Nominally significant associations are shown by colour. *Significant after Bonferroni correction for multiple testing.

To understand whether these results are influenced by confounding factors, we conducted comprehensive subgroup and sensitivity analyses to identify potential confounders and assess risk of bias. We found that our results were robust when correcting for age at score assessment (Supplementary Fig. 2, Supplementary Fig. 3). Results were also consistent when using raw non-normalized or z-score normalized scores instead of gene-level min-max normalization (Supplementary Fig. 4), supporting our choice of normalization while allowing for better clinical interpretation. Next, we investigated the influence of variant type: 94/648 (14.5%) of individuals in the study cohort had variants of unknown significance (VUS), but excluding these individuals did not impact the results. Likewise, no change was found when correcting for variant type including protein-truncating or missense variants (Supplementary Fig. 5). Lastly, we investigated the influence of epilepsy duration (time between seizure onset and score assessment) and epilepsy severity (including number of anti-seizure medications per year). Results were consistent for epilepsy duration, while the association for treatment with SCBs was not observed when correcting for epilepsy severity (Supplementary Fig. 6).

### Socioeconomic factors

Given that we found an association between socioeconomic factors and quality of life, we followed up by comparing specific score domains in detail. Three associations were nominally significant in the main analysis: Foster care or home placement, parent’s highest educational status, and annual household income (Fig. 1). Foster care or home placement had a nominally significant association with worse adaptive function in the main analysis (*β* = −0.084, 95% CI: [−0.164, −0.003], *p* = 0.043) but showed no difference at the specific score domain level. Overall, effect estimates were nominally significant but small. Nonetheless, they were consistent across domains and represented real-world differences: The strongest positive correlations of parents’ highest educational status were with child physical health (4 items, e.g. “had enough energy to participate in daily routines and activities”, *r* = 0.091, *p* = 0.033) and social interaction (7 items, e.g. “enjoyed the social experiences of meal times”, *r* = 0.087, *p* = 0.043). Similarly, annual household income most strongly correlated with social interaction (*r* = 0.140, *p* = 0.001), independence (5 items, e.g. “helped to complete routine activities”, *r* = 0.110, *p* = 0.016), and physical health (*r* = 0.100, *p* = 0.019; Fig. 3). For example, the mean QI-Disability score was 63.2 (σ = 15.9, *n* = 54) in families where the highest educational attainment was high school and 69.6 (σ = 14.6, *n* = 326) for four-year degrees (two-sided Wilcoxon rank-sum test: *p* = 0.044). Meanwhile, the mean QI-Disability score ranged from 64.8 (σ = 17.8, n = 64) for families with income in the US median income bracket ($36,000 - $50,000) to 71.6 (σ = 14.0, n = 117) for families with very high income (over $161,000 annual income; two-sided Wilcoxon rank-sum test: *p* = 0.030). Thus, we found consistent and domain-specific evidence of healthcare disparities associated with quality of life in individuals with NDD and epilepsy.

**Figure 3.**
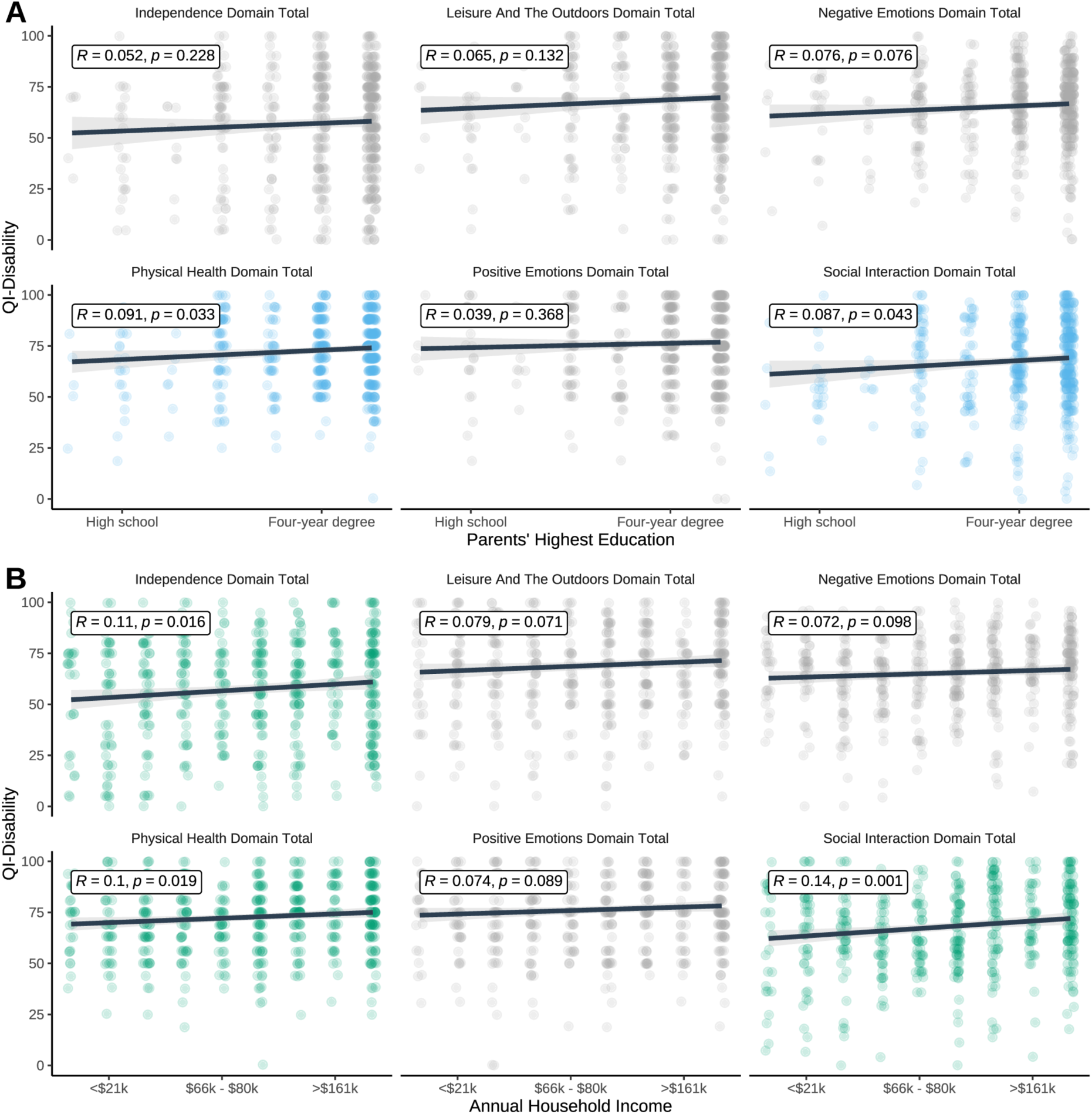
Association between parental education and annual income and the quality of life of children with genetic epilepsies. **(A)** Correlation between parental education and QI-Disability score. **(B)** Correlation between annual household income and QI-Disability score. Both exposures were associated with higher scores on the physical health and social interaction domains of the QI-Disability score. Linear regression with standard error shown as shaded contour and Pearson’s correlation coefficient. Nominally significant associations are shown by colour.

### Treatment-related factors

Having established the effect of socioeconomic factors on quality of life in NDD with epilepsy, we next examined treatment-related and thus potentially modifiable exposures. Hospitalisation was strongly associated with both lower quality of life and lower adaptive functioning in the main analysis. We found that this effect correlated with the frequency of hospitalisations (Vineland-3: *r* = −0.21, *p* < 0.001, QI-Disability: *r* = −0.18, *p* < 0.001, Fig. 4A; Fig. 4B). This finding also represented real-world differences: For example, individuals who were only hospitalized once had a mean Vineland-3 score of 57.2 (σ = 21.5, *n* = 266) while individuals who were hospitalized ≥5 times had a mean Vineland-3 score of 39.5 (σ = 17.6, *n* = 26; two-sided Wilcoxon rank-sum test: *p* < 0.001). For comparison, raw score changes of 1 to 3 points were considered clinically meaningful to the majority of caregivers in a study of Dravet syndrome, an NDD with epilepsy.^14^ This effect was independently seen as a correlation between quality of life and days spent in neonatal intensive care (NICU; *r* = −0.41, *p* = 0.025, *n =* 31, Fig. 4C). The associations with hospitalisations were profound across all domains of the QI-Disability score, especially for leisure and the outdoors (5 items, e.g. “enjoyed moving their body”, *r* = −0.16, *p* < 0.001; Fig. 4D). Similarly, almost all domain scores of adaptive functioning decreased with more frequent hospitalisations, most strongly for gross motor skills (43 items, e.g. “sits unsupported for at least 1 minute”, *r* = −0.16, *p* < 0.001; Fig. 4E), likely representing deconditioning.

**Figure 4.**
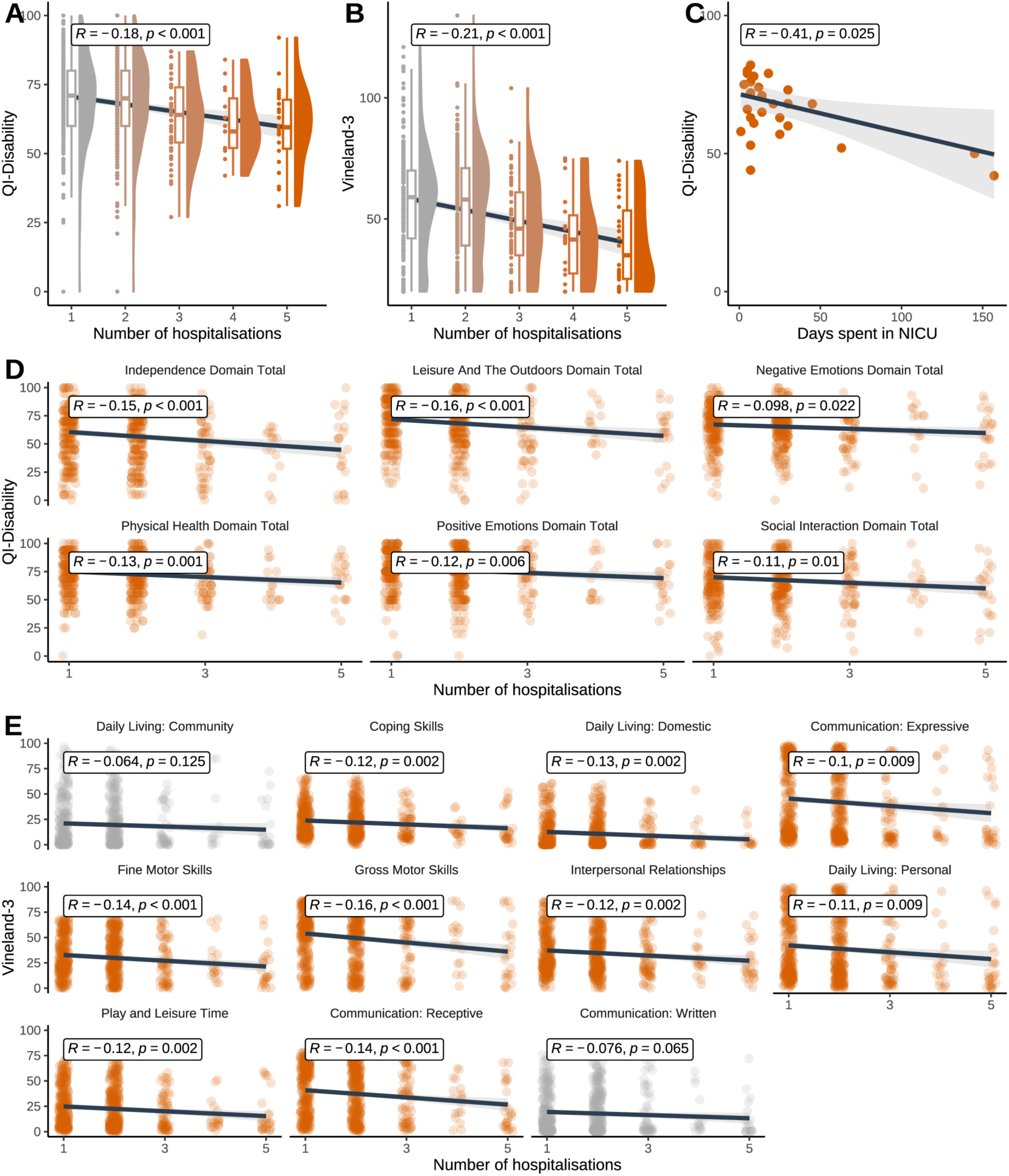
Association between hospitalisation and the quality of life and adaptive function of children with genetic epilepsies. (A-B) The negative effects of hospitalisations increase with the number of admissions. **(C)** More days spent in a neonatal intensive care unit (NICU) were linked to lower quality of life but not lower adaptive function. **(D)** Hospitalisations were associated with broad impairment on all domains of the QI-Disability score. **(E)** Hospitalisations were associated with broad impairment across most domains of the Vineland-3 score. Linear regression with standard error shown as shaded contour and Pearson’s correlation coefficient. Nominally significant associations are shown by colour.

Given this strong association, we suspected that reverse causality may influence our findings: Children who are sicker would be more likely to be admitted for inpatient care and may be at higher risk for readmission. Similarly, children who had epilepsy for longer, due to earlier onset or longer follow-up, may be more exposed to treatment-related factors. The association between hospitalisations and adaptive function was partially, but not fully, mediated by epilepsy duration (time between seizure onset and age at score assessment; average causal mediation effect, ACME: *β* = −0.001, 95% CI: [−0.001, −0.001], *p* = 0.043; average direct effect, ADE: *β* = −0.040, 95% CI: [−0.060, −0.020], *p* < 0.001; proportion mediated: 34%, 95% CI: [19, 55%]) and the number of ASMs received, as a surrogate parameter for epilepsy severity (ACME: *β* = −0.002, 95% CI: [−0.003, −0.001], *p* < 0.001; ADE: *β* = −0.040, 95% CI: [−0.060, - 0.020], *p* < 0.001; proportion mediated: 21%, 95% CI: [6, 45%]; Supplementary Fig. 7). This suggests that approximately one-third of the observed effect of hospitalisation on adaptive function is explained by the longer duration and greater burden of epilepsy treatment.

Among the two treatment-related factors modifying developmental outcomes identified in the main analysis, treatment with SCBs was most strongly associated with both lower quality of life and lower adaptive functioning (Fig. 1). Given prior literature on gene-specific treatment patterns and responses^33–35^, we hypothesized that this association may be related to gene- or disease-specific confounding factors and thus compared scores distributions for individuals with the six most common (*n* ≥ 30) genetic etiologies in the study cohort (*SCN2A*, *n* = 66; *STXBP1*, *n* = 60; *SLC6A1*, *n* = 55; *SYNGAP1*, *n* = 42; *PPP2R5D*, *n* = 41; 16p11.2 deletion, *n* = 33). Indeed, we did observe gene-level effects: Individuals with *STXBP1* variants who were treated with SCBs had a mean Vineland-3 score of 51.6 (σ = 17.8, *n* = 19) while individuals with *STXBP1* variants who were not treated with SCBs had a mean Vineland-3 score of 58.5 (σ = 17.6, *n* = 23; treatment data not available for the remaining patients; two-sided Wilcoxon rank-sum test: *p* = 0.017; Fig. 5A). Again, this difference in score range is considered meaningful to caregivers in another NDD with epilepsy.^14^ Similarly, individuals with *STXBP1* variants who were treated with SCBs had a mean QI-Disability score of 58.1 (σ = 21.2, *n* = 27) while individuals with *STXBP1* variants who were not treated with SCBs had a mean QI-Disability score of 71.7 (σ = 15.2, *n* = 33; two-sided Wilcoxon rank-sum test: *p* = 0.007). Quality of life was also lower in individuals with *SCN2A* variants treated with SCBs (mean score 58.2, σ = 14.6, *n =* 51 vs. mean score 67.2, σ = 15.0, *n =* 15; two-sided Wilcoxon rank-sum test: *p* = 0.040; Fig. 5B). Domain-specific associations were seen for leisure and the outdoors, negative emotions (7 items, e.g. “showed aggression”), and positive emotions [4 items (e.g., “showed happiness through body language”); Fig. 5C].

**Figure 5.**
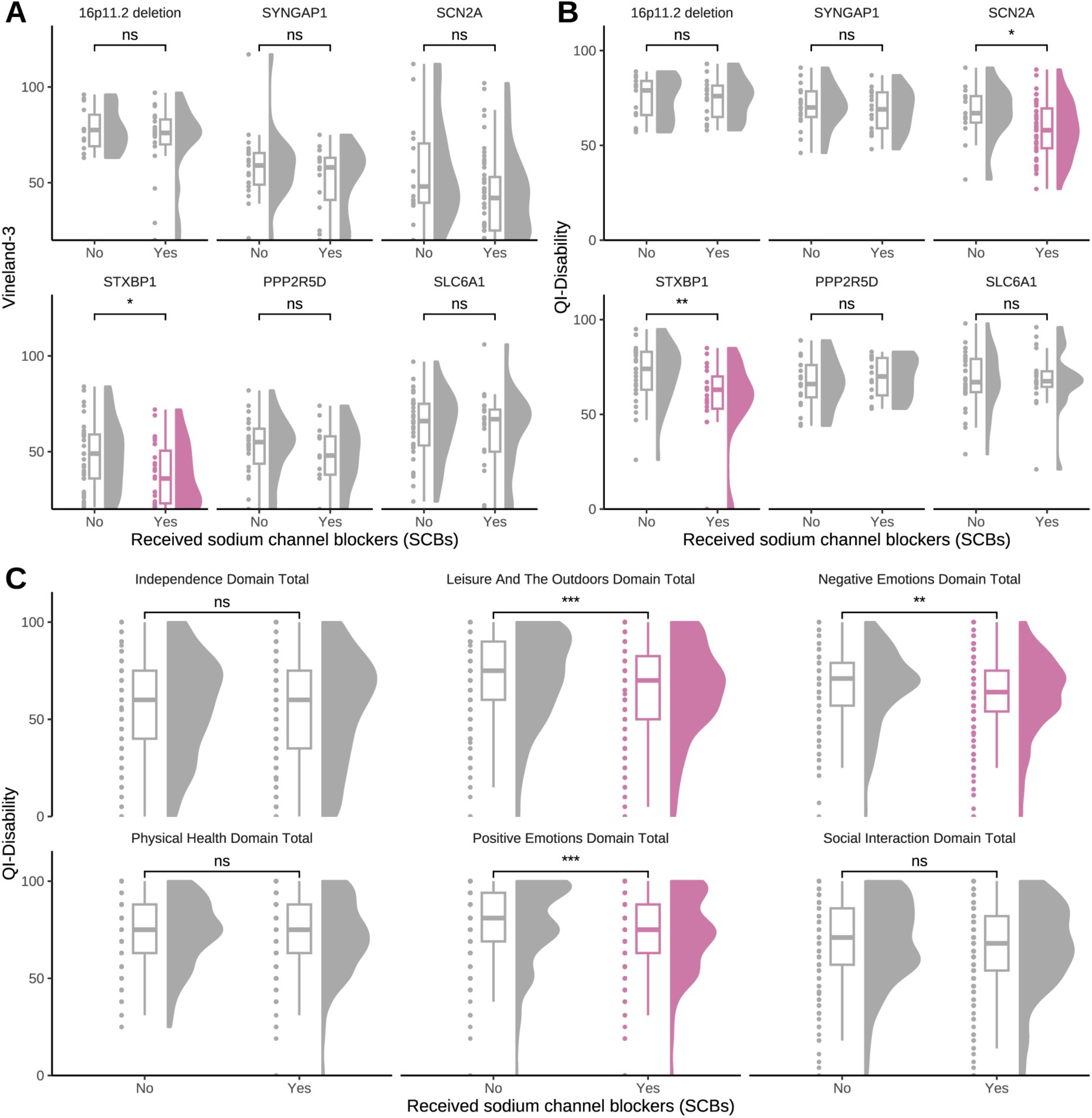
Association between treatment with sodium channel blockers and quality of life and adaptive function of children with genetic epilepsies. (A-B) The association with sodium channel blockers is primarily observed in individuals with variants in *SCN2A* or *STXBP1*. The six most common genetic etiologies in the cohort are shown. **(C)** Treatment with sodium channel blockers was associated with lower quality of life in the domains for leisure and the outdoors, negative emotions, and positive emotions. There were no domain-specific exposure effects on the Vineland-3 score. Two-sided Wilcoxon rank-sum test. Significance: *p < 0.05, **p < 0.01, ***p<0.001, ****p<0.0001. Nominally significant associations are shown by colour.

Given this strong association, we suspected that the same reverse causality identified for hospitalisations may also apply here: Children who had epilepsy for longer or had more treatment-resistant seizures may have received more and different ASMs including SCBs. Indeed, we found that the association between treatment with SCBs and adaptive function was partially, but not fully, mediated by epilepsy duration (ACME: *β* = −0.001, 95% CI: [−0.001, - 0.001], *p* < 0.001; ADE: *β* = −0.040, 95% CI: [−0.080, −0.040], *p* < 0.05; proportion mediated: 54%, 95% CI: [33, 97%]) and the number of anti-seizure medications (ACME: *β* = −0.006, 95% CI: [−0.002, −0.010], *p* < 0.01; ADE: *β* = −0.060, 95% CI: [−0.110, −0.020], *p* < 0.01; proportion mediated: 69%, 95% CI: [27, 100%]; Supplementary Fig. 7). Thus, the association between treatment with SCBs and adaptive function can be largely explained by both the longer duration and the greater burden of epilepsy treatment, similar to the effect of hospitalisations.

Based on the gene-level differences observed above (Fig. 5A-B), we also investigated if there are gene-specific mediation effects in the six most common (*n* ≥ 30) genetic etiologies in the study cohort. This hypothesis was based on the same prior literature on gene-specific treatment patterns and responses^33–35^, but formalizes the comparison of gene-level subgroups presented above (Fig. 5A-B) by further accounting for covariates and indirect effects. We found three examples: In *SCN2A*, mediation by treatment with SCBs was observed (ACME: *β* = −0.006, 95% CI: [−0.099, −0.021], *p* < 0.001; ADE: *β* = −0.240, 95% CI: [−0.300, −0.170], *p* < 0.001; proportion mediated: 7%, 95% CI: [2, 15%]). In *STXBP1*, there were only direct effects between the genetic aetiology and development (β = −0.150, 95% CI: [−0.220, −0.080], p < 0.001) and treatment with SCBs and development (*β* = −0.090, 95% CI: [−0.129, −0.050], *p* < 0.001), respectively. In *SLC6A1*, there was only a direct effect of treatment with SCBs on development (*β* = −0.090, 95% CI: [−0.129, −0.049], *p* < 0.001; Supplementary Fig. 8). Thus, the association of sodium channel blockers with developmental outcomes is both subject to gene-specific effects and linked to epilepsy duration and polytherapy burden.

Given this link to polytherapy burden, we wondered if this effect was specific to SCBs and therefore investigated another commonly used class of ASMs, GABAergic medication including benzodiazepines, barbiturates, stiripentol, tiagabine and vigabatrin. Treatment with GABAergic ASMs was correlated with even lower quality of life (*β* = −0.064, 95% CI: [−0.095, −0.032], *p* < 0.001) and adaptive function (*β* = −0.119, 95% CI: [−0.159, −0.078], *p* < 0.001; Supplementary Fig. 9A). Interestingly, other gene-specific subgroups could be identified, with the largest difference in adaptive function seen for individuals with *SYNGAP1* variants (Supplementary Fig. 9B), which may partially represent indication bias as it reflects gene-specific treatment patterns in practice.^36^ Treatment with GABAergic ASMs was associated with much lower scores across almost all domains of the QI-Disability and Vineland-3 scores when compared to exposure with sodium channel blockers (Supplementary Fig. 10). Together, these findings are in line with the role of gene-specific associations and polytherapy outlined above.

### Secondary analysis and replication

We replicated our main analysis using the SCQ Lifetime score, a caregiver-reported screening tool for ASD which measures behaviour related to social interaction, language development, and repetitive and restricted behaviours throughout the individual’s lifetime. This secondary analysis was done to reduce potential bias from using a single instrument to measure adaptive function (Vineland-3). Overall, results were variable depending on the screening threshold chosen from previous sensitivity analyses.^31,32^ We found that socioeconomic factors were negatively associated (parents’ highest educational status: *β* = −0.036, 95% CI: [−0.063, −0.009], *p* = 0.010; annual household income: *β* = −0.023, 95% CI: [−0.044, −0.003], *p* = 0.025) while foster care or home placement (*β* = 0.216, 95% CI: [0.006, 0.426], *p* = 0.044) and hospitalisations (*β* = 0.082, 95% CI: [0.033, 0.131], *p* = 0.001) were positively associated with being screened at increased likelihood for ASD (Fig. 6A). For example, the effect of hospitalisations translated to a ∼10% increased chance of being screened at increased likelihood for ASD (Odds Ratio, OR = 1.09, 95% CI: [1.03, 1.14], *p* = 0.001). Additionally, there was a nominally significant association between iron supplementation during pregnancy and being less likely to be screened at increased likelihood for ASD (*β* = −0.216, 95% CI: [−0.408, −0.025], *p* = 0.028). Exposure associations across the questions composing the SCQ Lifetime score were limited by multiple testing, but primarily demonstrated the same broad associations seen previously for hospitalisations. Thus, our secondary analysis found the same associations and, more importantly, the same effect directions observed in the main analysis except for treatment with SCBs. This suggests that the identified environmental influences generalize to broader aspects of development in NDD.

**Figure 6.**
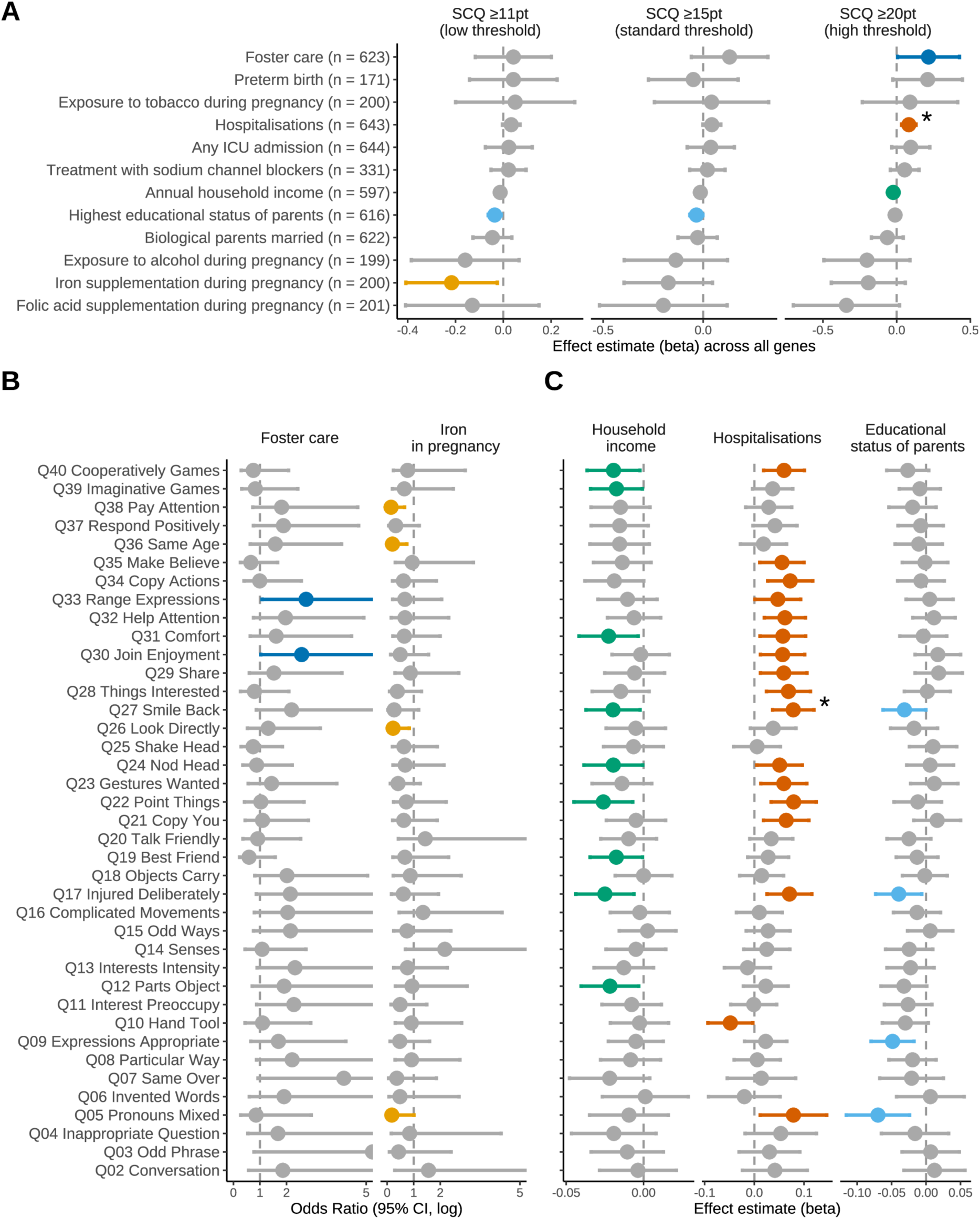
Replication of exposure associations with the Social Communication Questionnaire (SCQ) Lifetime. **(A)** Forest plot of the effect estimate (beta) of exposure across three commonly used screening thresholds. Multivariable logistic regression with sex, ethnicity, and age at seizure onset as additional covariates. For each variable, the number of individuals for which data was available is shown in brackets. **(B)** Exposure associations with questions on the SCQ Lifetime across binary exposures. Fisher’s exact test. **(C)** Exposure associations with questions on the SCQ Lifetime across continuous exposures. Note that Q01 is not shown as it is the initial classifier question for non-verbal children. Univariate logistic regression. Nominally significant associations are shown by colour. *Significant after Bonferroni correction for multiple testing.

So far, we have found that the associations in our main and secondary analysis are broadly consistent and that potentially modifiable treatment-related factors (hospitalisations, treatment with SCBs) are partially mediated by the duration and severity of epilepsy. Therefore, we hypothesized that some of these associations may be disease-specific for epilepsy. We repeated the main and secondary analyses in individuals with NDD without epilepsy (*n =* 322) and including a broader group of individuals with NDD where epilepsy status was unknown (*n =* 1598; Fig. 1). We found a nominally significant association between any ICU admission and Vineland-3 score (*β* = 0.129, 95% CI: [0.003, 0.256], *p* = 0.047) and QI-Disability score (*β* = 0.130, 95% CI: [0.025, 0.235], *p* = 0.016). However, none of the significant associations from the main analysis were seen in this cohort. In the replication of the secondary analysis, the associations and effect directions for foster care or home placement, hospitalisations, annual household income, parents’ educational status, and iron supplementation during pregnancy could be replicated (Supplementary Fig. 11). This suggests that some environmental factors may have gene-, disease-, or treatment-specific effects pending further validation.

### Missing heterogeneity explained in genetic epilepsy

Previously, we have found associations for parent’s educational status, annual household income, foster care or home placement, hospitalisations, treatment with SCBs, and iron supplementation during pregnancy with any of the three developmental outcome measures – representing six out of twelve pre-specified hypotheses. The effect size and statistical significance of each association partially varied across subgroups, sensitivity analyses, and score thresholds. However, this naturally raises the question how much all associations together can explain differences in developmental outcomes across NDD. We investigated this by fitting two models for each score: a base model with genetic aetiology, sex, race, and age at seizure onset as covariates; and a full model with all associations as additional covariates. Environmental exposures explained significantly more variance in developmental outcomes (Vineland-3; Akaike information criterion, Λ1_AIC_ = −15.84, ANOVA *p* = 0.003). Across all outcomes there was still a substantial amount of unexplained heterogeneity, ranging from 37.6% to 57.5% (Figure 7A). This suggests that additional predictors of developmental outcomes remain to be identified.

**Figure 7.**
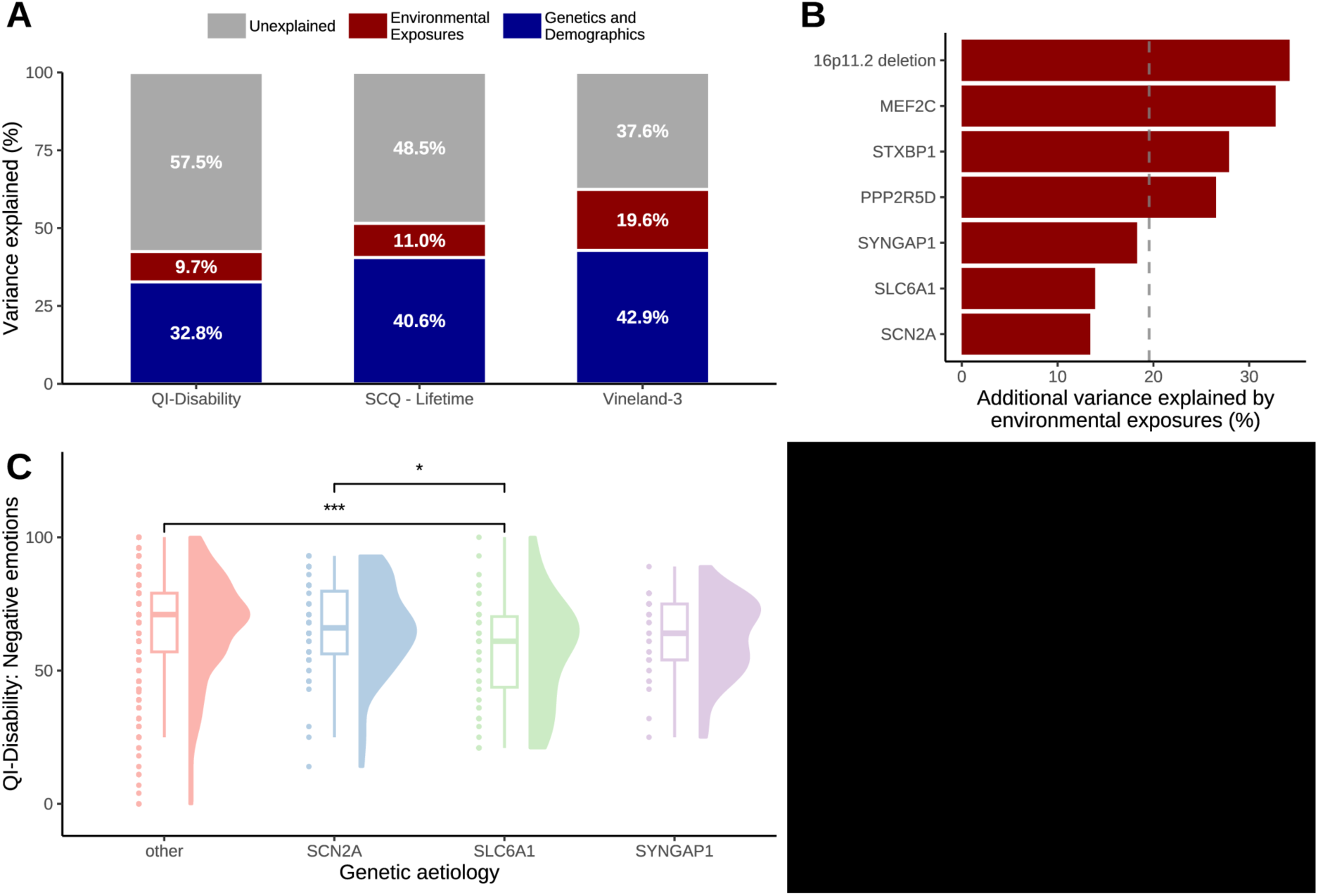
Missing heterogeneity explained in genetic epilepsy. **(A)** Bar chart of variance (*R*^2^) explained by a baseline model of genetic aetiology, sex, race, and age at seizure onset compared to the additional variance explained by environmental factors, where all significantly associated exposures were included as covariates. **(B)** Additional variance explained by the full model, shown separately for each of the most common genes in the study cohort. The dashed vertical line denotes the mean variance explained across the entire cohort (*R*^2^ = 19.6%). **(C)** Gene-specific differences in variance explained by environmental exposures may be related to strong gene-specific effects on behaviour, denoted here by the score on the ‘negative emotions’ domain of the QI-Disability score. Two-sided Wilcoxon rank-sum test. Significance: *p < 0.05, **p < 0.01, ***p<0.001, ****p<0.0001.

Interestingly, the additional variance explained by environmental exposures was different across genes (Figure 7B), again focusing on the most commonly observed genes in our study cohort (*n* ≥ 30). The lowest variances explained were seen for *SCN2A* (*R*^2^ = 13.4%), *SLC6A1* (*R*^2^ = 13.9%), and *SYNGAP1* (*R*^2^ = 18.3%). We hypothesized that specific genetic etiologies may more strongly determine developmental outcomes. Individuals with *SCN2A* or *SLC6A1* variants may show behavioural abnormalities including aggression^33,37^, which would be expected to heavily impact the caregiver-reported outcome measures. This was reflected by gene-specific lower scores on the ‘negative emotions’ domain of the QI-Disability (Fig. 7C). Together with the gene-specific mediation of treatment-related factors outlined above, this gene-specific variance suggests a varying degree of susceptibility to environmental factors.

## Discussion

Here, we provide first-time quantitative evidence that potentially modifiable environmental factors are associated with developmental outcomes including quality of life. Although specific genetic epilepsies have been studied in isolation^13,14,38^, we assessed a broad spectrum of 93 NDDs to provide comprehensive real-world evidence from the largest such cohort to-date. This approach directly highlights the unmet medical needs of this vulnerable population.

Treatment-related factors including hospitalisations and treatment with ASMs had broad associations with quality of life and adaptive function across most domains, showed a monotonic and dose-dependent relationship across the number of hospitalizations, and had convergent validity across scores with consistent signals found in our replication analysis. Thus, we consider these findings to be most credible and broadly applicable. The general impact of hospitalizations has been well-established in the general paediatric population and caregiver perspectives^22,39^, while we demonstrate the same effect in NDDs with epilepsy, highlight potential disease-specific confounders, and provide a quantitative effect estimate to inform risk management. Here, reverse causality and confounding by indication play a role: Children who are sicker are more likely to be admitted for inpatient care and are at higher risk for readmission.^40,41^ Appropriately, the association for hospitalisations remained only nominally significant after correcting for disease severity in sensitivity analyses, and the association for treatment with SCBs was not observed anymore. In early-onset seizures, SCBs are preferentially used for severe epilepsies where previous treatments have already failed which represents potential confounding by indication.^42–44^ This is further supported by the comparable signal seen for GABAergic ASMs. In epilepsy, disease severity is also correlated with age at seizure onset as most catastrophic genetic epilepsies manifest early in life^4,45^, which we corrected for by including age at seizure onset in our models.

However, our sensitivity and mediation analyses suggest that epilepsy duration and severity do not fully account for the observed treatment-related differences in developmental outcomes. This raises the possibility that these treatment-related factors may be potentially modifiable. For example, the risk of inappropriate hospitalisations or their sequelae may be reduced by: *(i)* guidelines on the prehospital treatment of seizures including avoidance of excessive medication^46^; *(ii)* education and seizure action plans to inform emergency management by caregivers and teachers^47^; *(iii)* improved medical treatment including newer anti-seizure medications^48^; *(iv)* continuation of developmental therapy during hospital stays and rehabilitation programs aimed at preventing deconditioning.^49,50^ Treatment with SCBs was primarily associated with polytherapy (i.e., greater burden of epilepsy treatment), which may be reduced by: *(i)* rationalizing polytherapy and minimizing drug-drug interactions^51,52^; *(ii)* use of newer ASMs, not necessarily because of higher efficacy – although cenobamate may represent an exception – but primarily due to their improved tolerability and safety.^53–55^ In turn, optimizing medical treatment may also reduce the risk of hospitalisations.^56^ Our findings suggests that, together, these ‘best practices’ aimed at safe treatment and early seizure control may protect the developmental outcomes of these often severely affected individuals.

Treatment-related effects of ASMs including SCBs were dependent on the underlying genetic aetiology. Partially, this association can also be related to the reverse causality outlined above as disorders associated with variants in genes encoding ion channels or synaptic proteins manifest early in life and can be particularly severe.^57,58^ This complex interplay may also explain why SCBs have been suspected of inducing epileptic spasms^59^, though this effect was not observed in epileptic spasms related to *STXBP1* variants.^34^ In *SCN2A*-related disorders, the differential effects of SCBs we observed may be related to functional mechanisms: missense variants may lead to gain-of-function (GOF) resulting in more severe early-infantile epilepsies that are amenable to treatment with SCBs, but they may also lead to loss-of-function (LOF) with later-onset epilepsies that lack response to SCBs.^33^ Similarly, the follow-up analysis of GABAergic ASMs revealed known treatment-related patterns in *SYNGAP1*.^36^ Interestingly, accounting for variant type (missense variants versus protein-truncating variants) strengthened the association for SCBs in our sensitivity analyses which further suggests variant-specific effects. In other genetic etiologies, e.g. *SLC6A1*, we found no interaction. These gene- and variant-specific effects have been investigated in cohort studies but have not previously been quantified while adjusting for confounders.^33,34,37^ Fully disentangling these effects and establishing causality will require large and prospective natural history studies. However, our findings further support the importance of genetically informed care for precision medicine.

Socioeconomic factors including parent’s education and annual household income were positively associated with quality of life, and provide first-time quantitative effect estimates to measure these healthcare disparities. While effect estimates were small, they were overall meaningful with a mean difference of ∼7 points on the 100-point scale or a change by a third of the standard deviation, above the minimally clinically important difference (4.83 points).^60^ These findings were further supported by the good re-test reliability, responsiveness, and robustness to seizure frequency of the QI-Disability.^60–62^ Together, these findings represent consistent evidence of healthcare disparities associated with quality of life in individuals with NDD and epilepsy. This aligns with prior evidence: children with epilepsy were more likely to live in lower-income households, were more likely to live in poorer living conditions, and may not receive adequate healthcare.^63–65^ Importantly, these prior findings were made in national population-based studies across all epilepsies. Here, we demonstrate that the same disparities also extend to individuals with NDD and epilepsy. We did not find the same disparities in individuals without epilepsy, which may suggest that seizure- or treatment-related factors contribute to these inequities. Reducing these healthcare disparities motivates strategic research initiatives, public health policy, and involvement of patient advocacy groups.^66^

Together, the six socioeconomic and treatment-related factors we identified better explain the differences in developmental outcomes than genetic and demographic information alone. Gene-environment interactions have previously been investigated to explain heterogeneity in the general population^67,68^ and have long been suspected to play a role in NDDs^69,70^, but have remained largely unexplored compared to genetic mechanisms. Here, we provide an estimate for the additional variance explained: In total, our models may explain one to two thirds of the variance in developmental outcomes. Together with the modifying factors and domain-specific associations outlined above, this information on developmental trajectories may inform natural history studies and guide clinical trial design on non-seizure related outcomes.^71,72^ Interestingly, the variance explained differed substantially across genetic etiologies. This may be explained by the wide range of phenotypic heterogeneity observed for each disorder.^33,35,73^ Further, we hypothesized that strong behavioural symptoms may profoundly impact quality of life and adaptive function measures and found evidence of gene-specific effects on aggression. Together, these findings further support genetically informed care. However, we also found a substantial amount of unexplained variance across developmental outcomes and quality of life, which suggests that additional predictors – across both genome and exposome – remain to be identified. This ‘missing heterogeneity’ motivates further data collection in natural history studies and the identification of oligogenic, polygenic and epigenetic influences.^74–76^

In summary, this study represents an initial systematic characterization of the environmental modifiers of developmental outcomes in genetic NDDs with epilepsy. We identified several directly treatment-relevant factors which collectively reframe genetic NDDs as genome-informed yet exposure-sensitive disorders with actionable opportunities to improve developmental trajectories.

### Limitations

This retrospective caregiver-reported study is limited to an observational design. The results of our causal mediation analyses should be considered as rough and exploratory estimates given the risk of unmeasured confounding and non-linear relationships between our predictors. Caregiver-reported measures may be subject to potential misclassification from recall bias and are not fully verified by clinicians. Additionally, the cohort was biased towards high-income white individuals and exclusively recruited from the United States, limiting generalizability of our findings to other settings. Almost half of individuals in the comparison group had at least one seizure without a formal diagnosis of epilepsy, which further demonstrates how common this comorbidity is across NDDs. However, this potential bias did not affect our main analyses, which were conducted within-group. Further, we restricted our study to a set of pre-specified hypotheses to reduce bias and increase transparency of our findings. Future studies on larger prospective cohorts may relax this approach to discover additional predictors, moving towards a comprehensive view of all environmental exposures (‘exposome’).^77,78^ In our study, some expected associations with developmental outcomes – such as smoking or alcohol use during pregnancy or the protective effect of folic acid supplementation – were not directly observed. This is most likely related to limited power compared to the large and well-designed population studies that previously demonstrated these effects. Alternatively, their impact may be less prominent in genetic epilepsies compared to the associations identified here. Other negative findings, such as the lack of an association between socioeconomic factors and adaptive functioning, are interesting by themselves as they had not previously been explored in NDDs. Lastly, correlation does not imply causation and the predictors of developmental outcomes we presented represent a complex interplay that cannot be reduced to single culprits: Many hospitalisations are necessary and helpful, and SCBs remain a generally effective and safe component of medical epilepsy management.

## Acknowledgements

We are grateful to all of the families at the participating Simons Searchlight sites as well as the Simons Searchlight Consortium, formerly the Simons VIP Consortium. We appreciate obtaining access to genetic and phenotypic data on SFARI Base.

## Supplementary material

Pending full publication.

